# Phase 1, randomized, crossover study comparing intravenous GTX-104 to oral nimodipine in healthy human subjects

**DOI:** 10.1101/2025.04.06.25325334

**Authors:** S. George Kottayil, Amresh Kumar, Carrie D’Andrea, Prashant Kohli, James Longstreth, R. Loch Macdonald

## Abstract

Enterally-administered nimodipine is the only approved drug formulation available in the United States for treatment of patients with aneurysmal subarachnoid hemorrhage. Intravenous nimodipine is available in other countries but it contains a high concentration of ethanol that is irritating to the vasculature, can alter the effects of other medications, impair neurological assessments and is potentially harmful to the liver. We developed a sterile aqueous solution of nimodipine solubilized in polysorbate 80 micelles (GTX-104) that circumvents these problems. GTX-104 has been administered to 168 healthy human volunteers in 2 studies. We report the second study here, a phase 1, single center, randomized, 2-period cross over study that assessed the pharmacokinetics of GTX-104 and oral nimodipine capsules, which is the reference standard, in 58 healthy human volunteers. GTX-104 was administered for 72 hours as a continuous infusion of 0.15 mg/hour with a 30 minute bolus infusion of 4 mg every 4 hours. Nimodipine capsules were administered orally at a dose of 60 mg every 4 hours for 72 hours. The maximum plasma concentrations after the first dose of each formulation were similar (GTX-104: 63 ng/mL, n=57 versus nimodipine capsules: 69 ng/mL, n=56, ratio and 90% confidence interval [CI] of geometric means: 92% [90% CI: 82-104%]). The areas under the concentration-time curves on the 3rd day at steady state also were the same (GTX-104: 497 ng*h/mL, n=55 versus nimodipine capsules: 495 ng*h/mL, n=56, ratio and 90% CI of geometric means: 106% [90% CI: 99-114%]). The secondary pharmacokinetic parameters (daily maximum concentration at steady-state and time to maximum concentration) were also similar for the 2 formulations. The variability in PK parameters was less for GTX-104 compared to oral nimodipine. The average oral bioavailability for nimodipine capsules was 7%. These results enabled a Phase 3 safety study of GTX-104 in humans with aneurysmal subarachnoid hemorrhage.

## Introduction

Subarachnoid hemorrhage (SAH) is a type of hemorrhagic stroke that is usually caused by a ruptured intracranial saccular aneurysm [1]. While the incidence is only about 6 per 100,000 population per year, the average age of those afflicted is almost 2 decades younger than for ischemic stroke, and the mortality approaches 50%. Outcomes for aneurysmal SAH (aSAH) have improved over time, although the only treatments shown to be effective in adequate, well-controlled clinical trials are nimodipine and repair of the ruptured aneurysm by coiling rather than clipping [2].

Formulations of nimodipine approved for use in the US, Canada and some other countries include tablets, capsules and a solution, all administered orally while intravenous (IV) nimodipine is available in some other countries [3–5]. There are drawbacks with oral nimodipine including variable and low bioavailability (F), interference of absorption with feeding and high first-pass metabolism. Numerous medications induce or inhibit hepatic cytochrome enzymes that metabolize nimodipine, leading to reduced or increased nimodipine plasma concentrations. Furthermore, it is difficult to administer the capsules or tablets to patients who cannot swallow. The oral solution causes diarrhea [3,6]. As a result of these problems, peak plasma concentrations are highly variable and can lead to unpredictable hypotension, which can be dangerous for patients with aSAH. Although clinicians respond to hypotension by holding or reducing the dose of nimodipine or stopping it altogether, dose reductions are associated with increased risk of delayed cerebral ischemia and poor outcome [7–10].

Intravenous nimodipine could avoid these problems. Compared to solid dose forms, the IV formulation that is approved in the European Union, United Kingdom, China and some other countries, provides high F (approaching 100%) and more stable plasma concentrations and therefore potentially easier control of blood pressure (BP) [11,12].

However, nimodipine is practically insoluble in water and the existing IV nimodipine formulation contains 23.7% (v/v) ethanol and 17% (v/v) polyethylene glycol 400. Ethanol causes liver toxicity, irritation of the blood vessel at the infusion site and impairs assessment of neurological function [13,14]. This report describes an IV formulation of nimodipine (GTX-104) that avoids these problems.

## Materials and methods

### GTX-104

GTX-104 is a sterile, aqueous-based, clear, colorless to light yellow solution containing nimodipine (2 mg/mL). Inactive ingredients include ultrapure polysorbate 80 (USP), dehydrated alcohol (USP) and water for injection (USP). GTX-104 is diluted in commonly used IV solutions to obtain a solution for infusion containing nimodipine, 0.08 mg/mL. This dosing solution is composed of dispersed micelles of polysorbate 80 containing nimodipine and about 1.26% w/v alcohol (Fig 1). All of the excipients are generally recognized as safe.

**Fig 1.**
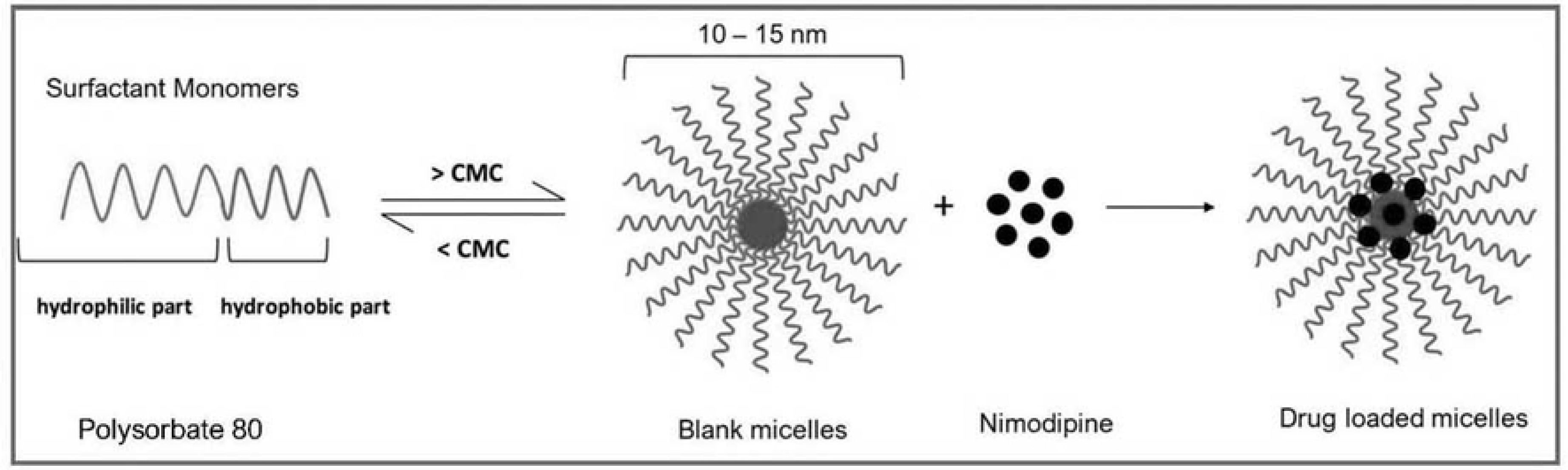
GTX-104 drug delivery technology.

### First in human study (GTX-104-001)

GTX-104 was administered to 104 healthy adult human volunteers in a 4-part, single center, randomized, dose-escalation and crossover study to assess the safety, tolerability, pharmacokinetics (PK) and absolute F of GTX-104 compared to nimodipine capsules. Overall, GTX-104 was well tolerated. There was one treatment emergent adverse event (TEAE, infusion site extravasation with GTX-104) and no serious adverse events (SAEs). Population PK modeling from this GTX-104-001 study served as the basis for the dose regimen used in the study described herein.

### Rationale, objectives and endpoints

The rationale for this study was to generate PK bridging data between IV and oral nimodipine capsules and to serve as a precursor to a GTX-104 Phase 3 study in patients with aSAH. The primary objective was to evaluate the relative F of IV GTX-104 versus nimodipine capsules at steady state in healthy human adults. The secondary objective was to compare safety and tolerability of these 2 formulations.

The primary study endpoints were maximum drug plasma concentration for the first dose on Day 1 (C_max Day 1 0-4h_) and area under the drug concentration-time curve (AUC) for 0 to 24 hours (h) on Day 3 (AUC_Day 3 0-24h_). Day 3 reflects PK steady state given the approximately 8-9 h elimination half life of nimodipine. Secondary PK endpoints were C_max_ on Day 3 for all 6 doses given that day (C_max Day 3 0-24h_), total and apparent total body clearance from plasma and absolute F.

Safety endpoints included the incidence of TEAEs and SAEs, their severity and relationship to study drug and change from baseline to the end of the study in vital signs, physical examination, clinical laboratory evaluations (hematology, clinical chemistry and urinalysis) and 12-lead electrocardiogram (ECG).

This study adhered to consensus ethical principles from the Declaration of Helsinki and the Council for International Organizations of Medical Sciences International Ethical Guidelines. It followed applicable International Conference for Harmonization and Good Clinical Practice and Laboratory Guidelines. All subjects gave informed consent in writing. Subjects were free to withdraw from the study at any time for any reason, without prejudice to their medical care.

### Study design

This was a Phase-1, single-center, randomized, 2-period crossover study in healthy, nonsmoking, normotensive male and female subjects between 18 and 55 years of age.

There was no blinding. Subjects had to be in good physical health, have normal cardiac conduction and function and have a body mass index between 18 and 32 mg/kg^2^. Subjects abstained from all medications except for the nimodipine formulations during the study.

The inclusion and exclusion criteria were selected to ensure subjects were healthy and that they did not have gastrointestinal or cardiovascular diseases that would compromise safety or PK data from the study (Table 1).

**Table 1.**
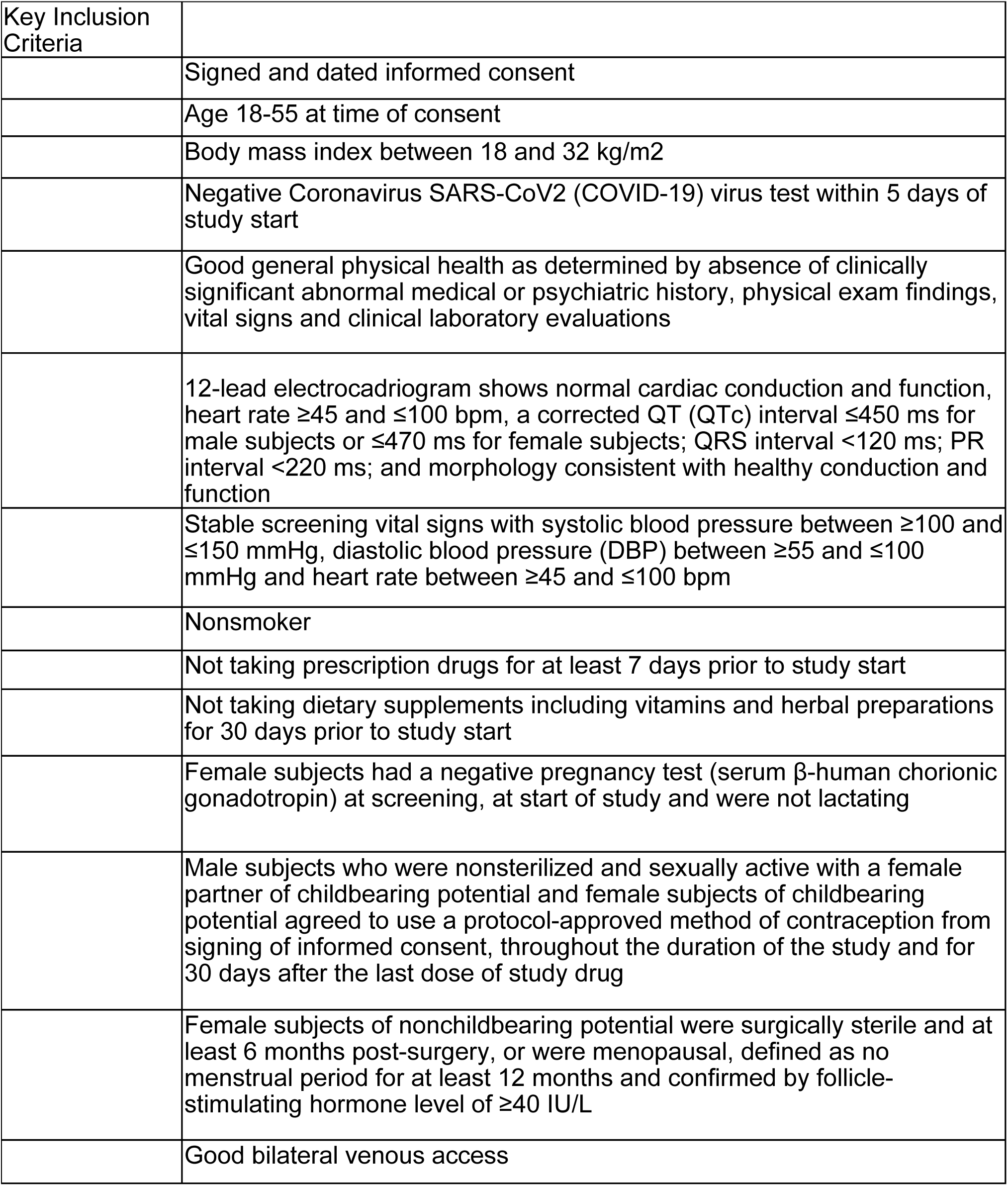
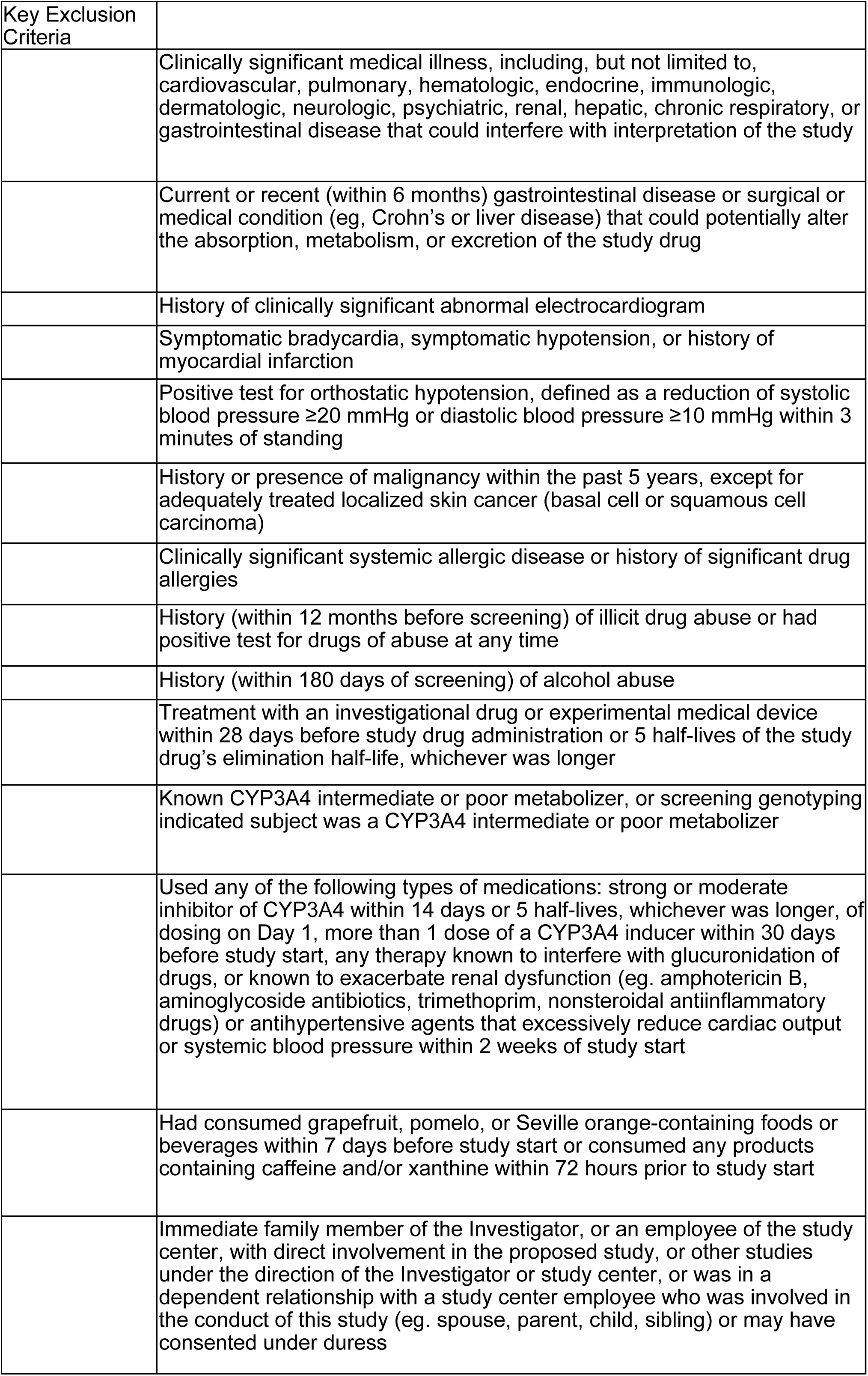
Key Inclusion and Exclusion Criteria.

On the day prior to starting drug dosing (Day -1), subjects were admitted to the clinical research unit where they remained domiciled for the duration of the study (Fig 3). They were randomly assigned by a computer generated schedule in a 1:1 ratio to receive GTX-104 first and nimodipine capsules second, or visa versa. The 2 treatment periods were separated by at least 96 h (Fig 3). On Days 1 to 4 of each treatment period, the study drug was administered at the same times each day (starting at 08:00 h on Day 1).

**Fig 2.**
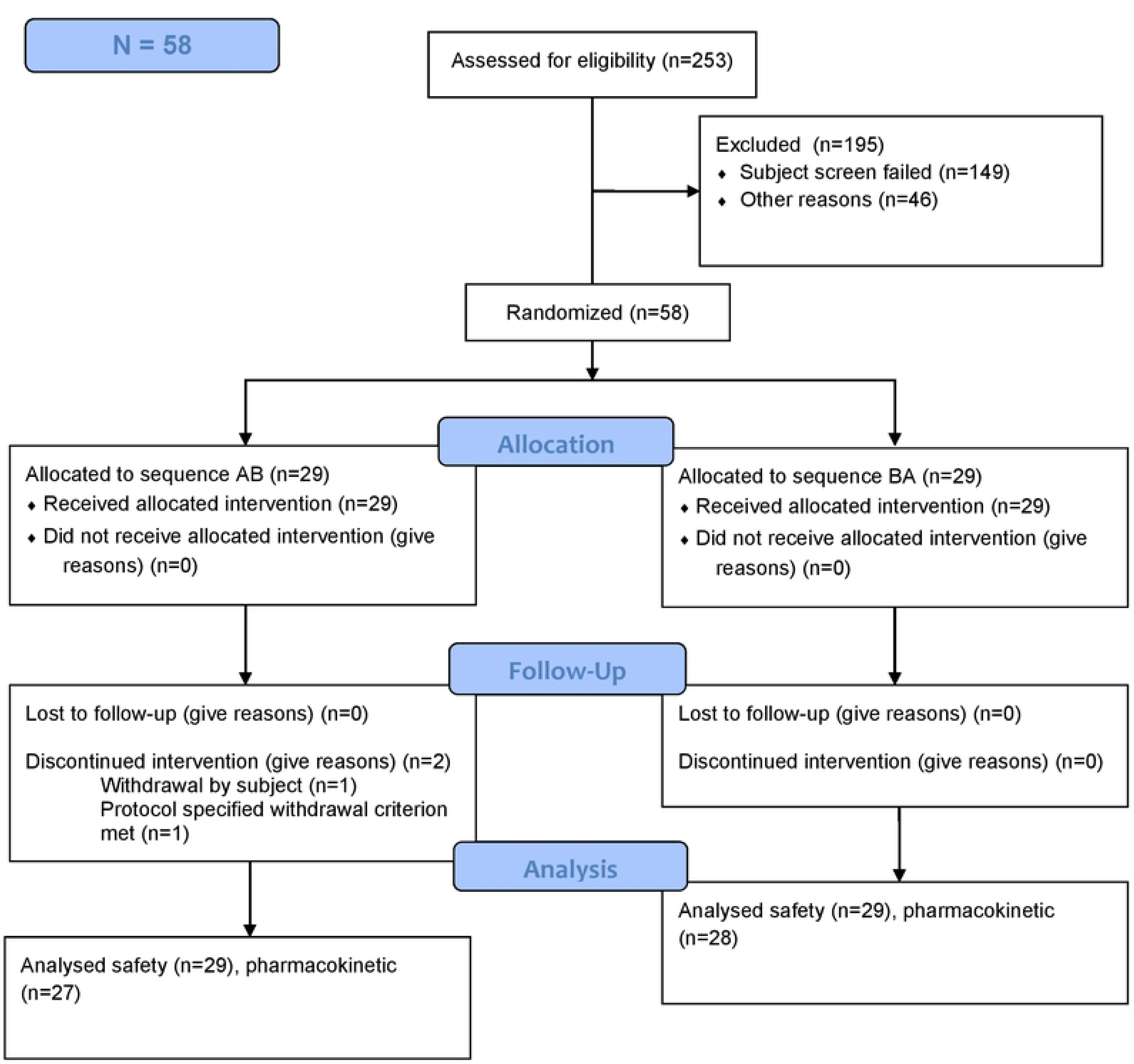
Study Profile Flowchart

**Fig 3.**
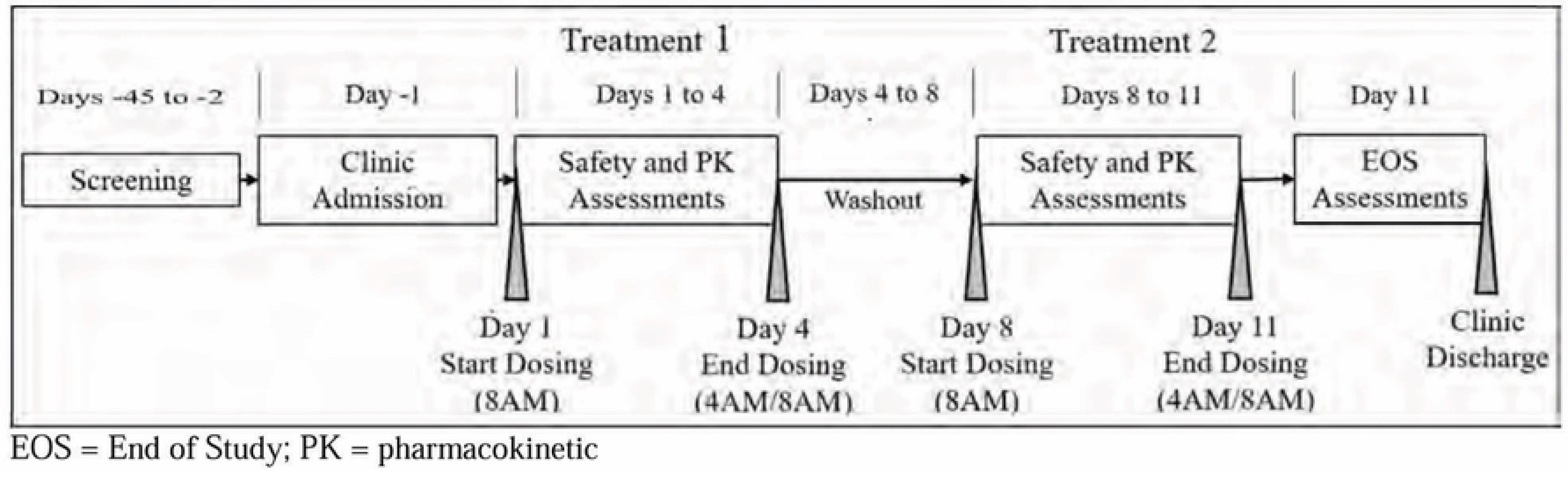
Study timeline.

Subjects fasted from 02:00 to 10:00 h so the doses at 04:00 and 08:00 h were administered in a fasting state. Standardized meals were provided at 10:00 h and then at approximately uniform times throughout the day.

The study was initiated August 26, 2021 and completed February 24, 2022. Altasciences Company, Inc. conducted the study, including screening and enrolling subjects, administering study drugs, monitoring subjects, obtaining clinical data and laboratory samples, performing bioanalytical laboratory tests and writing the clinical study report (Mount Royal, Quebec, Canada). All study source documents and the electronic case report forms are retained by Altasciences. The Sponsor was free to audit Altasciences at any time during the study. No amendments were made to the protocol once the study started.

### Drugs

GTX-104 was administered for 72 h as a 30-minute infusion of 4 mg every 4 h plus a continuous infusion of 0.15 mg/h. The GTX-104 dosing regimen was chosen to be safe and predicted by population PK modeling using data from the previous GTX-104-001 study to match the C_max_ and AUC for oral nimodipine capsules.

Nimodipine capsules (Nimotop®, reference formulation) were administered orally with 240 mL of water at a dose of 60 mg (two 30 mg capsules) every 4 h for 72 h.

### Assessments

Blood samples for nimodipine PK were taken around the first dose and on the third day. For the first dose, samples were taken at 08:00 h before the first dose and then 20, 30, 45, 60, 80 minutes and 2, 3 and 4 h after the dose. On the third day, the same scheduling was done around all 6 doses for that day, spanning from 48 to 72 h from the first dose. Blood pressure was taken prior to the first dose of study drug which was Day 1 and Day 8 for the second study drug. On Day 3 (and Day 10), it was taken 1 h after dosing for the 08:00, 12:00, 16:00 and 20:00 doses. Otherwise vital signs and BP were taken approximately every 12 h. Subjects were monitored continuously for AEs and SAEs.

Clinical laboratory evaluation was done at screening, Day −1 and at the end of the study. ECG was done at screening and at the end of the study.

Systolic and diastolic BP and heart rate changes were categorized based on whether they met one of 2 sets of criteria (Table 2).

**Table 2.** Definitions of Hypotension and Heart Rate Abnormaities.

| Blood Pressure | Category | Definition |
| --- | --- | --- |
|  | Hypotension 1 | Systolic BP <90 mmHg or diastolic BP <60 mmHg |
|  | Hypotension 2 | Systolic BP <90 mmHg or diastolic BP <60 mmHg or decrease in systolic or diastolic BP >10 mmHg compared with baseline |
| Heart Rate |  |  |
|  | Bradycardia 1 | heart rate <50 beats per minute |
|  | Bradycardia 2 | <50 or decrease >10 beats per minute |
|  | Tachycardia 1 | heart rate >100 beats per minute |
|  | Tachycardia 2 | heart rate >100 or increase >10 beats per minute |

Adverse events and medical monitoring were managed by Safe Harbor Pharmacovigilence, LLC, (Raleigh, North Carolina, USA). Adverse events were classified by system organ class and preferred term using the Medical Dictionary for Regulatory Activities, version 24.0 (Medrio database). The definitions and reporting of AE, TEAE, severity, seriousness and causality followed good clinical practice, the US government Federal Register (Code of Federal Regulations [CFR] Title 21, Part 312.32[a]) and International Conference on Harmonization guidelines. Laboratory and safety data were done in compliance with good laboratory practice. Relationship to study drug was determined by the study principle investigator based on clinical judgment, the Investigator Brochure and the nimodipine capsules prescribing information. Relationship was classified as unlikely (not related to study drug based on temporal relationship to study drug administration or for which other drugs, chemicals or underlying disease provide a plausible explanation) or possible/probable (related to study drug based on temporal proximity to study drug administration, possible or unable to be explained by other drugs, chemicals or underlying disease and for which information on drug withdrawal may be lacking, unclear or follows a reasonable response to drug withdrawal).

### Nimodipine assay

Blood samples for PK measurements were collected in K_2_EDTA vacutainers. As soon as possible following blood collection, samples were centrifuged for 10 minutes at 4°C and 1500g. The plasma obtained was separated into 2 polypropylene tubes. The samples were frozen at −20°C until assayed. The time from blood sample collection to plasma aliquot storage was not to exceed 90 minutes. Plasma samples were assayed for nimodipine using high-performance liquid chromatography with tandem mass spectrometry. The lower and upper limits of quantitation were 0.20 and 200.0 ng/mL, respectively.

### Sample size estimation

A model-based approach was used to estimate power and sample size. A population PK model was developed using data from the previous phase 1 GTX-104-001 study. This model included effects of food on the absorption of oral doses and a significant diurnal variation in F for the oral formulation. Simulated subjects (n=1000) were randomized to a 2-period (GTX-104 or nimodipine capsules) crossover. Day 1 (first dose) and Day 3 (steady state) simulated data were used to estimate the number of subjects needed for adequate power for the C_max Day 1 0-4h_ and AUC_Day 3 0-24h_ comparisons. The estimate for the within subject variance came from a mixed effects model that included fixed effects, treatment and a random effect for subject and period. Under the hypothesis that the true geometric mean ratio was 1 for GTX-104 compared to nimodipine capsules, 49 subjects were estimated to provide a power of 0.85 (C_max Day 1 0-4h_) and > 0.99 (AUC_Day 3 0-24h_), to assess relative F in a 2-period, 2-sequence crossover study.

A sample size of 60 subjects was planned in order to have at least 50 completed subjects.

### Statistical analysis

Safety was assessed on a safety population defined as all subjects who received at least 1 dose of study drug. The PK analysis was done on a PK population defined as all subjects who received at least one dose of study drug and had sufficient PK data to derive at least one PK parameter.

All demographics, clinical laboratory evaluations, vital signs, physical examinations, ECGs and other data were presented as listings and summary tables. The AEs and TEAEs were listings and summary tables by system organ class, preferred term, severity and relationship to study drug. All analysis, except PK, used SAS version 9.4 (Cary, North Carolina, USA).

### Pharmacokinetics

Missing nimodipine plasma concentration values were not imputed and were considered missing. Values below the lower limit of quantification were replaced with zero. PK data from the PK population for non-ratio variables and the analysis population for ratios were analyzed using Phoenix® WinNonlin® (Certara, Montreal, Quebec, Canada). T_max_ was analyzed descriptively. The natural logarithm transformed C_max Day 1 0-4h_, AUC_Day 3 0-24h_ and C_max Day 3 0-24h_ were used for all statistical inference and were analyzed using analysis of variance. Fixed factors included in the model were treatment received, the period at which it was given and the sequence in which each treatment was received. A random factor was added for the subject effect (nested within sequence). The treatment, sequence and period effects were evaluated at the 5% significance level. The 90% confidence intervals (CI) for the exponential of the difference in least-squares means (LS_means_) between each comparison was calculated for the log-transformed parameters (C_max Day 1 0-4h_, AUC_Day 3 0-24h_ and C_max Day 3 0-24h_). These values were back-transformed to provide the geometric ratios and 90% CI.

Relative F of GTX-104 and nimodipine capsules was calculated using the ratio of geometric LS_means_ with corresponding 90% CI calculated from the exponential of the difference between treatments for the natural log-transformed parameters C_max Day 1 0-4h_, AUC_Day 3 0-24hr_ and C_max Day 3 0-24h_.

## Results

Fifty-eight subjects were enrolled and randomized until January 2022 (Table 3; Fig 2). The PK population included all subjects who received at least one dose of the investigational product and had sufficient PK data to derive at least one PK parameter: 56 subjects in Period 1 and 55 subjects in Period 2.

**Table 3.** Demographic Characteristics.

|  |  | Sequence AB<br>(N=29) | Sequence BA<br>(N=29) | Overall<br>(N=58) |
| --- | --- | --- | --- | --- |
| Age (years) | Mean (SD) | 36.2 (6.94) | 38.3 (8.05) | 37.2 (7.52) |
|  | Median | 37.0 | 39.0 | 38.0 |
|  | Min, Max | 23, 48 | 22, 50 | 22, 50 |
| Gender [n(%)] | Male | 19 (65.5) | 19 (65.5) | 38 (65.5) |
|  | Female | 10 (34.5) | 10 (34.5) | 20 (34.5) |
| Ethnicity [n(%)] | Hispanic/Latino | 9 (31.0) | 11 (37.9) | 20 (34.5) |
|  | Not Hispanic/Latino | 20 (69.0) | 18 (62.1) | 38 (65.5) |
| Race [n(%)] | Asian | 1 (3.4) | 0 | 1 (1.7) |
|  | Black or African American | 1 (3.4) | 0 | 1 (1.7) |
|  | White | 25 (86.2) | 29 (100.0) | 54 (93.1) |
|  | Other | 2 (6.9) | 0 | 2 (3.4) |
| Weight (kg) | Mean (SD) | 81.52 (17.026) | 74.49 (10.260) | 78.01 (14.377) |
|  | Median | 84.10 | 74.80 | 79.55 |
|  | Min, Max | 46.7, 108.5 | 54.5, 93.7 | 46.7, 108.5 |
| Height (cm) | Mean (SD) | 173.36 (10.395) | 169.26 (8.249) | 171.31 (9.528) |
|  | Median | 176.90 | 169.20 | 173.30 |
|  | Min, Max | 151.1, 189.6 | 157.0, 185.7 | 151.1, 189.6 |
| Body Mass Index<br>(kg/m<sup>2</sup>) | Mean (SD) | 26.8 (3.64) | 26.0 (2.92) | 26.4 (3.29) |
|  | Median | 28.0 | 26.0 | 27.0 |
|  | Min, Max | 19, 31 | 21, 31 | 19, 31 |

Abbreviations: Max = maximum; Min = minimum.

### Pharmacokinetics

The geometric mean C_max Day 1 0-4h_ for GTX-104 and nimodipine capsules were 63 ng/mL and 69 ng/mL (intra-subject coefficient of variation [CV]: 39%, Fig 4, Table 4). The ratio of geometric LS_means_ for C_max Day 1 0-4h_ was 92% (90% CI: 82-104%). The variability in C_max Day 1 0-4h_ was lower for GTX-104 compared to nimodipine capsules (geometric CV: 26% versus 55%, respectively).

**Fig 4.**
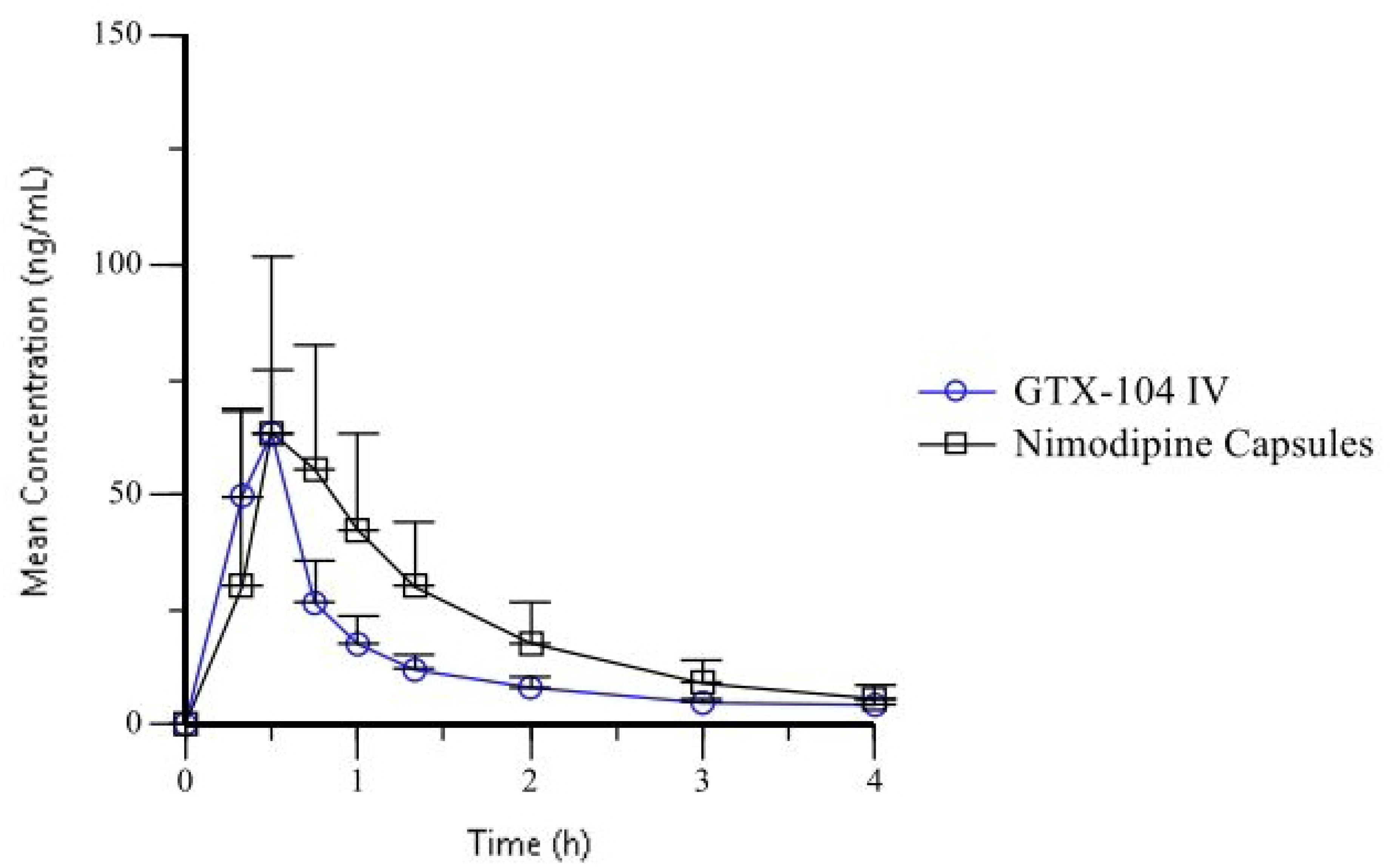
Mean (± standard deviation) Plasma Nimodipine Concentration after First Dose.

**Table 4.** Pharmacokinetic Parameters for Primary Endpoints.*

| Variable | GTX-104 | Nimodipine Capsules | Ratio (%; 90% CI) |
| --- | --- | --- | --- |
| $C_{\max}$ Day 1 0-4hr (geometric Least Squares mean, ng/mL) | 63.1 | 68.6 | 92 (82-104) |
| $AUC_{\text{Day 3 0-24hr}}$ (ng*h/mL) | 492 | 462 | 106 (99-114) |
| F (%; fraction of drug) | 100 | 7 |  |

The overall daily exposures on Day 3 (AUC_Day 3 0-24h_) were similar between GTX-104 and nimodipine capsules with geometric means of 492 ng*h/mL and 462 ng*h/mL, respectively (intra-subject CV: 22%, Fig 5). As with C_max Day 1 0-4h_, greater variability was observed for capsules compared to GTX-104 (geometric CV: 38% versus 16%, respectively). The ratio of geometric LS_means_ for the AUC_Day 3 0-24h_ was 106% (90% CI: 99-114%).

**Fig 5.**
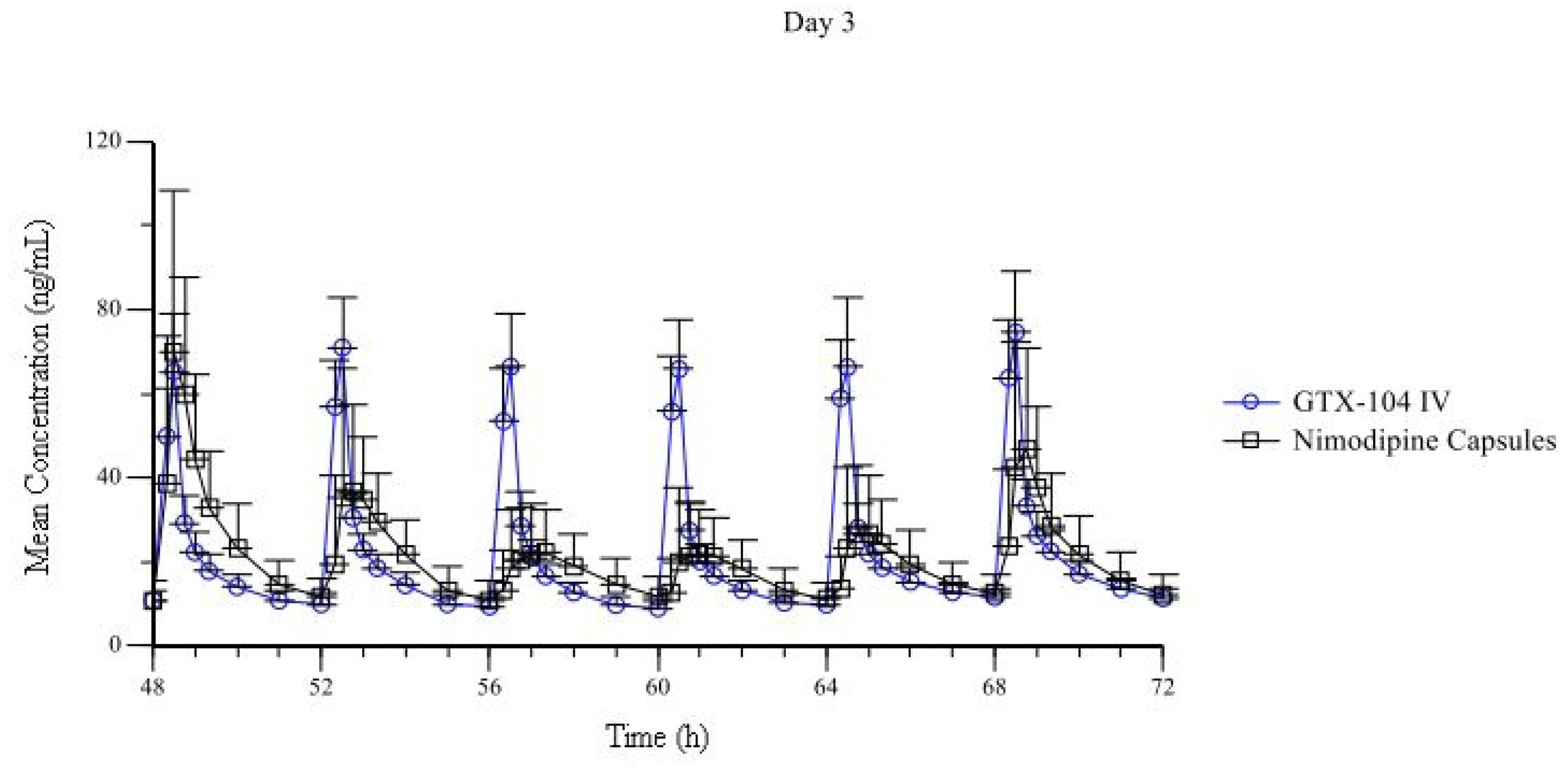
Mean (± standard deviation) Plasma Nimodipine Concentration after Each Dose on Day 3.

The geometric LS_mean_ C_max Day 3 0-24h_ for GTX-104 was 78 ng/mL compared to 85 ng/mL for nimodipine capsules; however, the variability was lower following GTX-104 administration (geometric CV: 17% versus 41% for nimodipine capsules, respectively). These maximum exposures for the 2 nimodipine treatments were similar at steady state on Day 3, with the ratio and 90% CI of the geometric LS_means_ for C_max Day 3 0-24h_ being 92% (90% CI: 85%-100%). The variability in C_max_ on Day 3 for GTX-104 was limited (geometric CV: 19-30%) over the 6 bolus doses, while C_max_ for nimodipine capsules showed greater variability (geometric CV: 46-61%).

The geometric mean apparent total body clearance on Day 3 for GTX-104 was 56 L/h (geometric CV: 15%) and for nimodipine capsules it was 772 L/h (geometric CV: 38%). Based on the ratio of nimodipine clearance during GTX-104 administration and apparent clearance during nimodipine capsule administration, the average oral bioavailability of nimodipine capsules was 7% (geometric mean, geometric CV: 32%).

Nimodipine plasma concentrations increased rapidly following both GTX-104 and nimodipine capsule administration, with median T_max_ values occurring 0.5 h after the first dose on Day 1 for both formulations (Table 5). On Day 3, median T_max_ was approximately 0.5 h for both treatments although greater variability in the range was observed following oral dosing (Table 5).

**Table 5.** T_max_ for first dose on Day 1 and for all doses on Day 3.

| Parameter | GTX-104 |  | Oral Nimodipine |  |
| --- | --- | --- | --- | --- |
|  | Median | Range | Median | Range |
| $T_{\max}$ Day 1 0-4 hr | 0.5 | 0.33-0.55 | 0.5 | 0.33-1.33 |
| $T_{\max}$ Day 3 0-24 hr | 0.5 | 0.50-0.57 | 0.62 | 0.33-2.00 |

### Adverse events

Overall, 44 out of the 58 subjects (76%) who received GTX-104 reported 122 TEAEs while 33 out of the 56 subjects (59%) who received nimodipine capsules reported 92 TEAEs (Table 6). Drug-related TEAEs were similar between groups (36/58 [62%] of GTX-104 and 31/56 [55%] of nimodipine capsule subjects). Most TEAEs were mild and considered related to study drug (GTX-104: 80%, nimodipine capsules: 87%). There were no serious AEs.

**Table 6.**
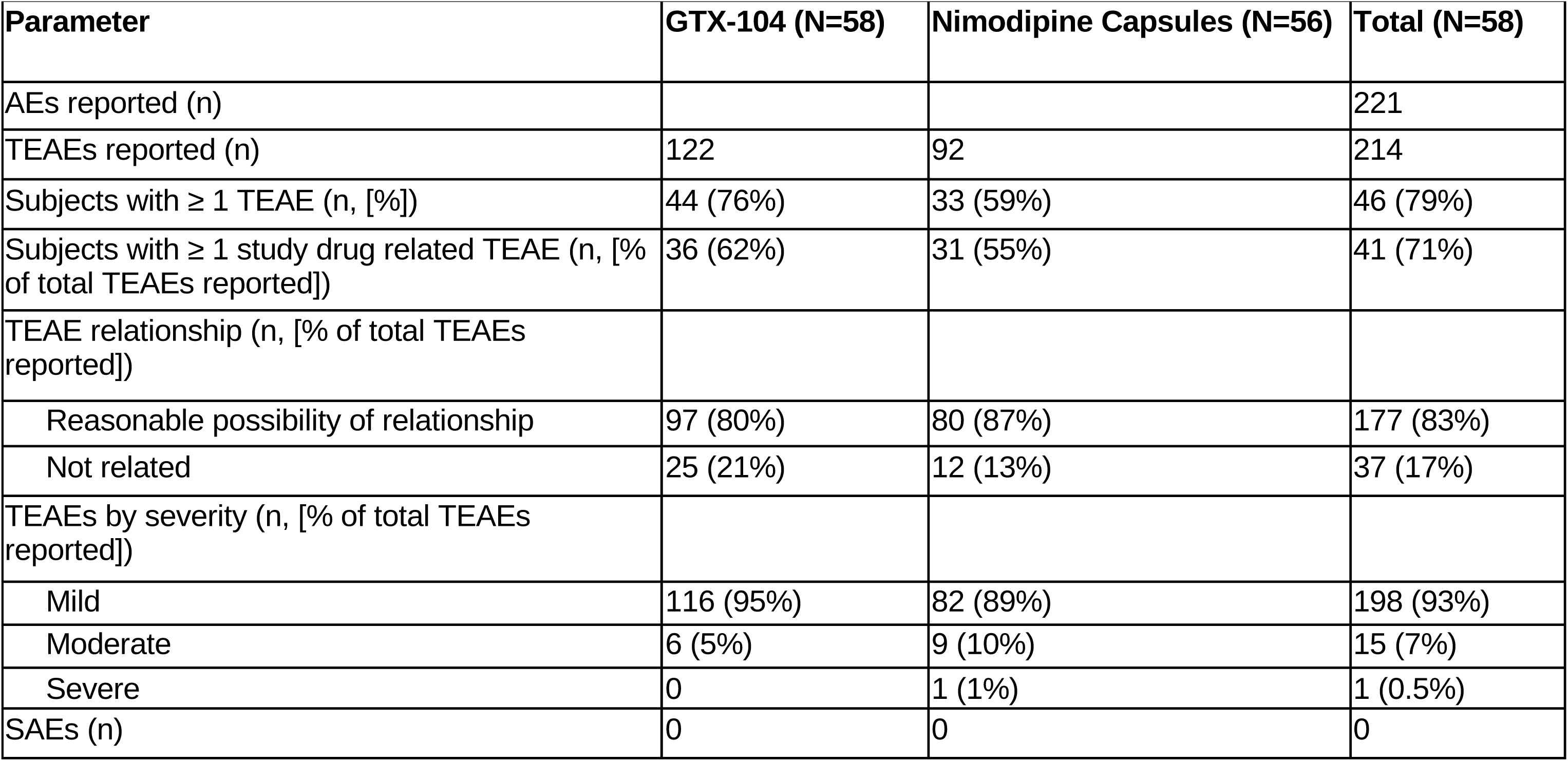
Overview of Adverse Events.

The most common system organ class affected was the nervous system, with 26 subjects (45%) reporting nervous system TEAEs following GTX-104 administration and 28 subjects (50%) following nimodipine capsule administration (Table 7). The most frequently reported TEAEs (reported by 2 or more subjects) were headache (GTX-104: 35%, nimodipine capsules: 36%), catheter site erythema (24%, 0%), somnolence (7%, 13%) and hot flushing (3%, 11%). Fewer gastrointestinal disorders were reported with GTX-104 than with nimodipine capsules (7% versus 16%, respectively) while more administration and sampling site events were reported with GTX-104 (41% versus 11%).

**Table 7:** Treatment Emergent Adverse Events by Medical Dictionary for Regulatory Activities Preferred Term and System Organ Class.

| System Organ Class | GTX-104 (N=58) | Nimodipine Capsules (N=56) | Total (N=58) |
| --- | --- | --- | --- |
| **Nervous system disorders (n [%])** | 26 (45%) | 28 (50%) | 34 (59%) |
| Headache (n [%]) | 21 (36%) | 20 (36%) | 25 (43%) |
| Somnolence (n [%]) | 5 (9%) | 7 (13%) | 10 (17%) |
| Dizziness (n [%]) | 4 (7%) | 3 (5%) | 7 (12%) |
| **General disorders and administration site conditions (n [%])** | 24 (41%) | 6 (11%) | 27 (47%) |
| Catheter site erythema (n [%]) | 15 (26%) | 0 | 15 (26%) |
| Catheter site pain (n [%]) | 3 (5%) | 2 (4%) | 5 (9%) |
| **Vascular disorders (n [%])** | 6 (10%) | 6 (11%) | 10 (17%) |
| Hot flush (n [%]) | 3 (5%) | 6 (11%) | 8 (14%) |

The severe TEAE occurred in a 30 year-old male with no medical history who was not taking any medications before entry into the study. On Day 1 of the first study drug cycle, the subject experienced syncope (reported term: fainted) following a catheter insertion attempt. This was before study drug (GTX-104) was administered. The event resolved immediately and was reported as an AE of syncope of severe intensity. During the second study drug cycle, the subject received 60 mg nimodipine capsules every 4 h between Day 8 and Day 11. The subject fainted at 14:15 on Day 11, approximately 2 h and 15 minutes after receiving the second daily dose and 1 h and 45 minutes prior to receiving the third daily dose of nimodipine capsules. The event resolved immediately and was reported as a TEAE of syncope of severe intensity. The Investigator considered the TEAE to be possibly related to drug administration. No concomitant medication was taken by the subject, and he completed the study per protocol.

Following administration of GTX-104, 38-41% (n=39) subjects experienced hypotension 2 (Tables 2 and 8). These changes were relatively consistent over the 4 boluses studied. The frequency of hypotension 2 following administration of oral nimodipine was 29-50% (n=39, Table 8). Overall, the proportion of subjects experiencing hypotension 1 was 33% (n=19) with GTX-104 and 45% (n=25) with nimodipine capsules. Approximately the same number of subjects in each group developed tachycardia 2 (heart rate >100 or >10 beats per minute above baseline). There were no clinically significant changes in laboratory tests or ECGs.

**Table 8.** Summary of Vital Sign Interpretation by Treatment.^a^.

|  | **Baseline**<br>**(N=56)** | **1 h**<br>**(N=56)** | **5 h**<br>**(N=56)** | **9 h**<br>**(N=56)** | **13 h**<br>**(N=56)** | **Overall**<br>**(N=56)** |
| --- | --- | --- | --- | --- | --- | --- |
| **Hypotension 1** |  |  |  |  |  |  |
| GTX-104 | 1 (2%) | 4 (7%) | 7 (13%) | 9 (16%) | 11 (20%) | 19 (34%) |
| Nimodipine capsules | 1 (2%) | 15 (27%) | 11 (20%) | 12 (21%) | 6 (11%) | 25 (45%) |
| **Hypotension 2** |  |  |  |  |  |  |
| GTX-104 | 1 (2%) | 21 (38%) | 22 (39%) | 22 (39%) | 23 (41%) | 39 (70%) |
| Nimodipine capsules | 1 (2%) | 28 (50.0) | 22 (39%) | 20 (36%) | 16 (29%) | 39 (70%) |
| **Bradycardia 1** |  |  |  |  |  |  |
| GTX-104 | 1 (2%) | 2 (4%) | 2 (4%) | 2 (4%) | 2 (4%) | 2 (4%) |
| Nimodipine capsules | 4 (7%) | 2 (4%) | 2 (4%) | 1 (2%) | 1 (2%) | 3 (5%) |
| **Bradycardia 2** |  |  |  |  |  |  |
| GTX-104 | 1 (2%) | 5 (9%) | 6 (11%) | 2 (4%) | 4 (7%) | 7 (13%) |
| Nimodipine capsules | 4 (7%) | 4 (7%) | 4 (7%) | 3 (5%) | 2 (4%) | 5 (9%) |
| **Tachycardia 1** |  |  |  |  |  |  |
| GTX-104 | 0 | 0 | 0 | 0 | 0 | 0 |
| Nimodipine capsules | 0 | 0 | 0 | 1 (2%) | 0 | 1 (2%) |
| **Tachycardia 2** |  |  |  |  |  |  |
| GTX-104 | 0 | 3 (5%) | 3 (5%) | 21 (38%) | 15 (27%) | 25 (45%) |
| Nimodipine capsules | 0 | 8 (14%) | 5 (9%) | 16 (29%) | 19 (34%) | 27 (48%) |

<sup>a</sup> Definitions of variables are in Table 2

## Discussion

Nimodipine is a dihydropyridine with a molecular weight of 418 g/mol that was developed in the 1970s as a vasodilator with specificity for the cerebral vasculature. It has and continues to be studied as a treatment for many diseases but currently has approval in North America for treatment of patients with aSAH. The formulations approved in the United States and Canada are administered enterally. Intravenous nimodipine is used in Europe, China and some other countries. The enteral formulations have limitations. The oral tablets and capsules cannot be swallowed by patients who are unconscious or intubated and methods to administer them through nasogastric tubes often result in inadequate dosing [3]. There is a risk of medication errors; fatalities have resulted from incorrectly injecting the liquid aspirated from capsules IV. An oral liquid formulation was developed to prevent this but it contains polyethylene glycol which causes diarrhea [3,6]. Bioavailability of oral dosage forms is poor. Only 2 reports of F in aSAH patients were found which reported a range of 3 to 28% in 36 patients [15,16]. Soppi, et al., were unable to detect nimodipine in the plasma of 5 of 8 aSAH patients despite receiving the drug enterally [11]. Nimodipine is metabolized by the CYP3A4 system and is thus subject to food effects in the intestines and to first pass effect in the liver, which are highly variable in patients with aSAH. Numerous medications induce or inhibit CYP3A4, leading to reduced or increased nimodipine plasma concentrations. This variability leads to side effects, principally hypotension, which is regarded as being detrimental to patients with aSAH.

If hypotension occurs, the dose of nimodipine is reduced. However, this may reduce its efficacy [7,10,17]. Ditz and colleagues retrospectively assessed the effect of nimodipine dose reduction in 205 patients during the high-risk period for cerebral vasospasm and delayed cerebral ischemia between 5 and 10 days following SAH [9]. Patients with reduction in nimodipine dosage had worse clinical and radiological grades and developed vasospasm and delayed cerebral ischemia significantly more often. They also were more likely to have unfavorable outcome.

An IV nimodipine formulation could circumvent several of these problems but one has never been approved in the United States. The currently available formulation contains high concentrations of alcohol and organic solvent (ethanol [23.7% v/v] and polyethylene glycol 400 [17% v/v]) that are not suitable for continuous IV infusion. It irritates blood vessels and has to be administered through a central venous catheter. The ethanol is associated with liver injury and it can hinder neurological assessments. It is administered by continuous IV infusion which does not replicate the sawtooth PK profile produced with the repeated dosing of nimodipine capsules. The importance of this PK pattern for efficacy is unknown, but in consultation with FDA scientists, a study design was adopted with the intent to demonstrate that an IV dosing regimen could produce a similar sawtooth pattern of exposure, including similar magnitudes of peak and total exposure, as oral capsules for the aSAH indication.

This report describes an IV formulation of nimodipine that does not have toxic or undesirable excipients. A dose regimen was selected to match the PK profile of nimodipine capsules. This IV formulation, GTX-104, was administered to try to replicate the PK of nimodipine capsules. It was shown that C_max Day 1 0-4h_ was similar following the first dose of both formulations with the ratio of geometric mean C_max Day 1 0-4h_ being 92% (90% CI: 82%-104%). The AUC_Day 3 0-24hr_ was measured on Day 3, which was taken to represent steady state. The cumulative exposures for the two nimodipine formulations were similar with the ratio of geometric means being 106% (90% CI: 99%-114%). The T_max_ for the first dose and for all the doses on Day 3 was also the same for GTX-104 and nimodipine capsules. The most notable other finding was that all PK parameters were less variable for GTX-104 compared to nimodipine capsules.

GTX-104 also was safe. Hypotension is the main clinically important side effect of nimodipine. Indeed, hypotension, defined as systolic BP <90 mmHg or diastolic BP < 60 mmHg, was less common with GTX-104 compared to nimodipine capsules. This was probably because there was less variability in C_max_ with GTX-104. Since unpredictable C_max_ is believed to cause hypotension with oral nimodipine, the more consistent C_max_ would be advantageous for patients with aSAH {Laursen1988}. It is possible that some instances of study drug effect on BP were not detected since scheduled BP measurements were 1 h after study drug administration. The other adverse events were mild and related to the injection site. The nimodipine capsule prescribing information does not reveal any other consistent, common side effects of nimodipine [18]. Headache is perhaps one such side effect although in the setting of aSAH it is essentially ubiquitous regardless of nimodipine use.

Bioavailability of nimodipine capsules was approximately 7%. Although there was essentially 100% bioavailability with GTX-104, there was less risk of hypotension. This is probably due to the high protein binding of nimodipine leaving little free drug to act on vascular L-type calcium channels.

There is great interest in the use of nimodipine as intraarterial rescue therapy for angiographic vasospasm in patients with aSAH [19,20]. Nimodipine also is injected into the ventricles of these patients for the same purpose [21,22]. These treatments have undergone some study in randomized clinical trials but additional ones are needed to determine their safety and efficacy. GTX-104 warrants study for these routes of administration, and potentially for other diseases [23].

## Conclusions

An IV formulation of nimodipine, GTX-104, that does not contain toxic excipients was developed. In this Phase 1 crossover design comparative study, GTX-104 and oral nimodipine capsules demonstrated largely similar results for the 2 primary endpoints (C_max Day 1 0-4 hr_ and AUC_Day 3, 0-24hr_). The secondary PK parameters of the daily C_max_ at steady-state and T_max_ values were also supportive of the PK comparability of the two formulations. The range of values observed for all PK parameters was wider for nimodipine capsules compared to GTX-104. The improved bioavailability, less frequent hypotension and more consistent PK suggest GTX-104 delivered IV has the potential to be a more convenient, efficient and controlled way to deliver nimodipine to patients with aSAH than currently available formulations. Moreover, GTX-104 can potentially improve the management of hypotension and vasospasm in SAH patients, thereby improving outcomes and potentially reducing mortality and/or long-term disability. A phase 3 randomized clinical trial was completed in January, 2025 and the results are not available yet [24].

## Data Availability

All relevant data are within the manuscript and its Supporting Information files

